# Evaluation of chest X-Ray with automated interpretation algorithms for mass tuberculosis screening in prisons

**DOI:** 10.1101/2021.12.29.21268238

**Authors:** Thiego Ramon Soares, Roberto Dias de Oliveira, Yiran E. Liu, Andrea da Silva Santos, Paulo Cesar Pereira dos Santos, Luma Ravena Soares Monte, Lissandra Maia de Oliveira, Chang Min Park, Eui Jin Hwang, Jason R. Andrews, Julio Croda

## Abstract

**Rationale:** The World Health Organization (WHO) recommends systematic tuberculosis (TB) screening in prisons. Evidence is lacking for accurate and scalable screening approaches in this setting.

**Objectives:** To assess the diagnostic accuracy of artificial intelligence-based chest x-ray interpretation algorithms for TB screening in prisons.

**Methods:** Prospective TB screening study in three prisons in Brazil from October 2017 to December 2019. We administered a standardized questionnaire, performed chest x-ray in a mobile unit, and collected sputum for confirmatory testing using Xpert MTB/RIF and culture. We evaluated x-ray images using three algorithms (CAD4TB version 6, LunitTB and qXR) and compared their diagnostic accuracy. We utilized multivariable logistic regression to assess the effect of demographic and clinical characteristics on algorithm accuracy. Finally, we investigated the relationship between abnormality scores and Xpert semi-quantitative results.

**Measurements and Main Results:** Among 2,075 incarcerated individuals, 259 (12.5%) had confirmed TB. All three algorithms performed similarly overall with AUCs of 0.87-0.91. At 90% sensitivity, only LunitTB and qXR met the WHO Target Product Profile requirements for a triage test, with specificity of 84% and 74%, respectively. All algorithms had variable performance by age, prior TB, smoking, and presence of TB symptoms. LunitTB was the most robust to this heterogeneity, but nonetheless failed to meet the TPP for individuals with previous TB. Abnormality scores of all three algorithms were significantly correlated with sputum bacillary load.

**Conclusions:** Automated x-ray interpretation algorithms can be an effective triage tool for TB screening in prisons. However, their specificity is insufficient in individuals with previous TB.

## INTRODUCTION

Globally, tuberculosis (TB) incidence in prisons is more than ten times higher than the general population (1). This disparity is especially alarming in South America, where TB cases in prisons have more than doubled since 2000 amid rising incarceration rates (1, 2). Several factors contribute to the elevated risk of TB in prisons, including overcrowding, poor ventilation, high rates of smoking, drug use, and limited access to medical care, leading to delays in TB diagnosis (2).

Interventions to address this growing burden are urgently needed, including improvements in case detection. In 2021, the World Health Organization (WHO) released updated guidelines on screening for TB, upgrading to a strong recommendation that systematic screening be conducted in prisons and penitentiary institutions (3). However, the recommendation is based on “very low certainty of evidence”, and guidance on specific means for screening in this setting is lacking. Moreover, correctional health systems are often underfunded and poorly equipped, and few prison systems in low- and middle-income countries perform systematic screening for TB despite the widely acknowledged high burden. Therefore, effective, cost-efficient screening approaches are needed to bring case-finding to scale in these settings. An important part of such approaches is a point-of-care triage test that can substantially reduce the number of people who need further testing.

Chest radiography is among the oldest tools for pulmonary TB screening and historically played a major role in TB control programs in high burden settings (4, 5). However, by the 1970s, concerns were raised about the accuracy, logistics and personnel requirements for mass radiography, leading the WHO to conclude in its 9^th^ expert committee report that “indiscriminate TB case finding by mobile mass radiography should be abandoned” (6).

Recently, there has been a resurgence in interest in the use of radiography as a screening tool for TB, leveraging recent advances in machine learning approaches to automate x-ray interpretation (7, 8). Clinic-based evaluations have demonstrated promising accuracy for several automated interpretation systems among individuals with TB symptoms. However, it is less clear how well these algorithms will perform for the purpose of active case finding, irrespective of symptoms, in incarcerated populations with high prevalence of smoking, drug use, and history of TB. To address this knowledge gap, we evaluated the performance of three deep learning-based x-ray interpretation algorithms in the context of mass screening for TB in three high burden prisons in Brazil.

## MATERIALS AND METHODS

### Study Design and Participants

We performed a cross-sectional study from October 2017 to December 2019 in three male prisons in Mato Grosso do Sul State, Brazil: Jair Ferreira de Carvalho Penitentiary (EPJFC), Campo Grande Penal Institute (IPCG), and Dourados State Penitentiary (PED). The prisons have a combined population of approximately 5,500 individuals. All incarcerated individuals in each prison were invited to participate in TB screening. Those who agreed to participate in the study provided written informed consent. The study was approved by the institutional review boards (IRBs) of the Federal University of Grande Dourados (UFGD) (#3.483.377) and Stanford University (#40285).

### Study Procedures

We outfitted a Volkswagen Constellation 24-240 truck with a 9.8 × 2.5-meter container, lead covering, an access ramp, an x-ray machine (Altus ST 543 HF, Sawae^®^), an x-ray scanner and digitizer (Agfa 15-X CR, Mortsel Belgium) and a separate room for sputum processing with two 4-module GeneXpert machines (Cepheid, Sunnyvale, USA). The mobile screening team consisted of a nurse, a laboratory technician, and an x-ray technician, with a physician available for consultation.

Study nurses administered a structured questionnaire to obtain demographic data, incarceration history, lifestyle factors, health history, and TB symptoms as recommended by the World Health Organization (WHO) (6). A spot sputum sample was collected from all participants who were able to produce sputum, with a target volume of at least 2 ml. Sputum samples were tested by Xpert MTB/RIF G4 (Cepheid, Sunnyvale, USA) and, if volume was sufficient, culture on Ogawa-Kudoh media. *M. tuberculosis* growth in cultures was confirmed by an immunochromatographic assay (TB Ag MPT64 Rapid Test, Standard Diagnostics, Seoul, South Korea).

A posterior-anterior chest x-ray was performed for all participants and then scanned and digitized. The images were electronically transferred for automated analysis by Computer-Aided Detection for TB version 6 (CAD4TBv6), developed by the Analysis Group at Radboud University Medical Center (Netherlands); Lunit INSIGHT CXR2 (hereinafter LunitTB) developed by the South Korean medical software company Lunit; and qXR, developed by Qure.ai in Mumbai, India. All information was recorded in Research Electronic Data Capture (REDCap^®^) (9, 10).

### Outcome Definitions and Analytic Approach

We defined TB cases as individuals with a positive Xpert MTB/RIF or culture growing *M. tuberculosis*. We defined controls as individuals who had sputum testing for Xpert MTB/RIF and no positive result by Xpert or culture. As a secondary analysis, we included as controls all individuals who were screened for TB and did not have a positive test, regardless of whether they were able to provide sputum. Individuals already undergoing treatment for TB were excluded from all analyses.

For CAD4TBv6, which provides a score range of 0 to 100, we used a positivity threshold score of _≥_60 through calibration with radiographic imaging data from a subset of participants with (n = 80) and without (n = 200) microbiologically confirmed TB. For LunitTB, which provides a score range of 0 to 1, we used a threshold of _≥_0.72 as specified by the manufacturer and identified through prior calibration (10) (11). For qXR (score range 0 to 1), we used a threshold of _≥_ 0.5 according to a previous study (11). We also evaluated the performance of each algorithm with the WHO’s Target Product Profile (12) (TPP) for a triage test by identifying the threshold that achieved 90% sensitivity and examining the corresponding specificity.

For each algorithm we calculated the sensitivity, specificity, positive predictive value (PPV), negative predictive value (NPV), and area under the receiver operating characteristic (ROC) curve (AUC). We calculated exact binomial confidence intervals (CIs) for sensitivity and specificity. We compared algorithm AUCs using DeLong’s test. For demographic and clinical characteristics, continuous variables were compared using the Mann-Whitney U test and categorical variables using the chi-square test. To assess the influence of demographic and clinical characteristics on algorithm performance, we conducted multivariable logistic regression controlling for age, race, drug use, smoking, previous TB, and presence of any TB symptoms (cough, fever, night sweat, weight loss, loss of appetite, tiredness, and chest pain). We report predicted marginal estimates of specificity for each characteristic at the WHO TPP threshold of 90% sensitivity. Finally, we investigated the relationship between Xpert semi-quantitative result and x-ray algorithm score among confirmed TB cases using Spearman’s rank correlation testing. Data were analyzed using SPSS version 25.0 and R version 4.0.3, including the pROC package (13).

## RESULTS

Between October 2017 and December 2019, we enrolled 7,081 participants across three male prisons in the Brazilian state of Mato Grosso do Sul. Sixty-six participants were excluded from further analyses as they were already under treatment for tuberculosis. Among the remainder, 2075 participants were able to produce valid sputum samples for Xpert and were included in the primary analysis (**Figure E1)**. Participants in the primary analysis had a median age of 33 years (IQR 28-40) **(Table 1)**. Compared with participants who did not produce a valid sputum, those who did had a higher prevalence of TB symptoms (73% vs 18%, p<0.001), smoking (73% vs 55%, p<0.001), illicit drug use (70% vs 54% p<0.001), and previous tuberculosis (14% vs 5%, p<0.001) **(Table E1)**.

**Table 1.** Sociodemographic characteristics and risk factors for TB among study participants, stratified by TB status as determined by sputum Xpert or culture.

| **Variables** | **Total**<br>**N=2075 (%)** | **TB cases**<br>**N=259 (%)** | **No TB**<br>**N=1816 (%)** | **P Value** |
| --- | --- | --- | --- | --- |
| Median age (IQR) | 33 (28, 40) | 33 (28, 39) | 33 (28, 40) | 0.497 |
| Prison Unit |  |  |  |  |
| PED | 889 (42.8) | 65 (25.1) | 824 (45.0) | <0.001 |
| EPJFC | 840 (40.5) | 144 (55.6) | 696 (38.3) |  |
| IPCG | 346 (16.9) | 50 (19.3) | 296 (16.3) |  |
| Race/ethnicity |  |  |  |  |
| Mixed | 1279 (61.6) | 158 (61.0) | 1121 (61.4) | 0.192 |
| White | 508 (24.5) | 56 (21.6) | 452 (24.9) |  |
| Black | 253 (12.2) | 38 (14.7) | 215 (11.8) |  |
| Indigenous | 33 (1.6) | 7 (2.7) | 26 (1.4) |  |
| Asian | 2 (0.1) | - | 2 (0.1) |  |
| <8 years of schooling | 1546 (74.5) | 198 (76.4) | 1348 (74.2) | 0.230 |
| Current smoker | 1520 (73.3) | 208 (80.3) | 1312 (72.2) | 0.006 |
| Illicit drug use over the last year | 1460 (70.4) | 203 (78.4) | 1257 (69.2) | 0.003 |
| Previously incarcerated | 1557 (75.0) | 214 (82.6) | 1343 (74.0) | 0.003 |
| BCG vaccinated | 1862 (89.7) | 223 (86.1) | 1639 (90.3) | 0.039 |
| Previous TB | 293 (14.1) | 55 (21.2) | 238 (13.1) | 0.000 |
| Report any WHO TB symptoms | 1512 (72.9) | 196 (75.7) | 1316 (72.5) | 0.277 |
| Report cough | 1255 (60.5) | 172 (66.4) | 1083 (59.6) | 0.037 |
| TB contact | 1565 (75.4) | 211 (81.5) | 1354 (74.6) | 0.160 |

**Abbreviations:** BCG: Bacillus of Calmette and Guérin; IQR: interquartile range; TB: tuberculosis; WHO:
World Health Organization, y: years.

During the screening period, 259 (12.5%) participants were diagnosed with pulmonary TB, of which 113 were diagnosed by sputum Xpert alone, 17 were diagnosed through sputum culture alone, and 129 had positive Xpert and culture tests. The prevalence of any TB symptom did not differ between TB cases and controls (76% vs 73%, p=0.277); however, cough was slightly more common among TB cases (66 vs 60%, p=0.037). Smoking, drug use, history of incarceration, and history of TB were significantly more prevalent among TB cases compared to non-TB cases (**Table 1**).

Among TB cases, 209 (80.7%) were classified as positive by CAD4TBv6, 207 (79.9%) by LunitTB, and 193 (74.5%) by qXR when using pre-defined thresholds (**Table 2)**. At 90% sensitivity, only LunitTB and qXR met the WHO’s Target Product Profile (TPP) with specificity of 83.7% (95% CI 72.4-87.3) and 74.2% (95% CI 60.2-81.3), respectively. At a 4% prevalence of TB, LunitTB had the highest PPV (18.7%), followed by qXR (12.7%) and CAD4TBv6 (9.0%). Receiver operating characteristic (ROC) curves for each algorithm are shown in **Figure 1**. Compared with CAD4TBv6 (AUC 0.877), LunitTB (AUC 0.912, p=0.0003) and qXR (AUC 0.901, p=0.0112) had higher AUCs, though AUC did not differ between LunitTB and qXR (p=0.17). In a secondary analysis of diagnostic accuracy in which we included the 4940 participants unable to provide sputum (total N=7081), AUCs did not differ substantially from the primary analysis, with LunitTB at 0.926, qXR at 0.920, and CAD4TBv6 at 0.904.

**Table 2.** Sensitivity, Specificity, Area Under the Curve (AUC), Positive Predictive Value (PPV) and Negative Predictive Value (NPV) of each algorithm at pre-defined thresholds or with thresholds adjusted to 90% sensitivity as specified by the WHO Target Product Profile minimum target.

| System | AUC (95% CI) | At pre-defined thresholds |  | At 90% sensitivity, 4% prevalence |  |  |
| --- | --- | --- | --- | --- | --- | --- |
|  |  | Sensitivity %<br>(95% CI) | Specificity %<br>(95% CI) | Specificity %<br>(95% CI) | PPV % | NPV % |
| CAD4v6 | 0.87 (0.85-0.90) | 80.7 (75.4-85.3) | 82.7 (80.8-84.4) | 62.3 (52.0-73.1) | 9.0 | 99.3 |
| LunitTB | 0.91 (0.88-0.93) | 79.9 (74.5-84.6) | 89.8 (88.3-91.2) | 83.7 (72.4-87.3) | 18.7 | 99.5 |
| qXR | 0.90 (0.87-0.92) | 74.5 (68.8-79.7) | 89.4 (87.9-90.8) | 74.2 (60.2-81.3) | 12.7 | 99.4 |

**Figure 1:**
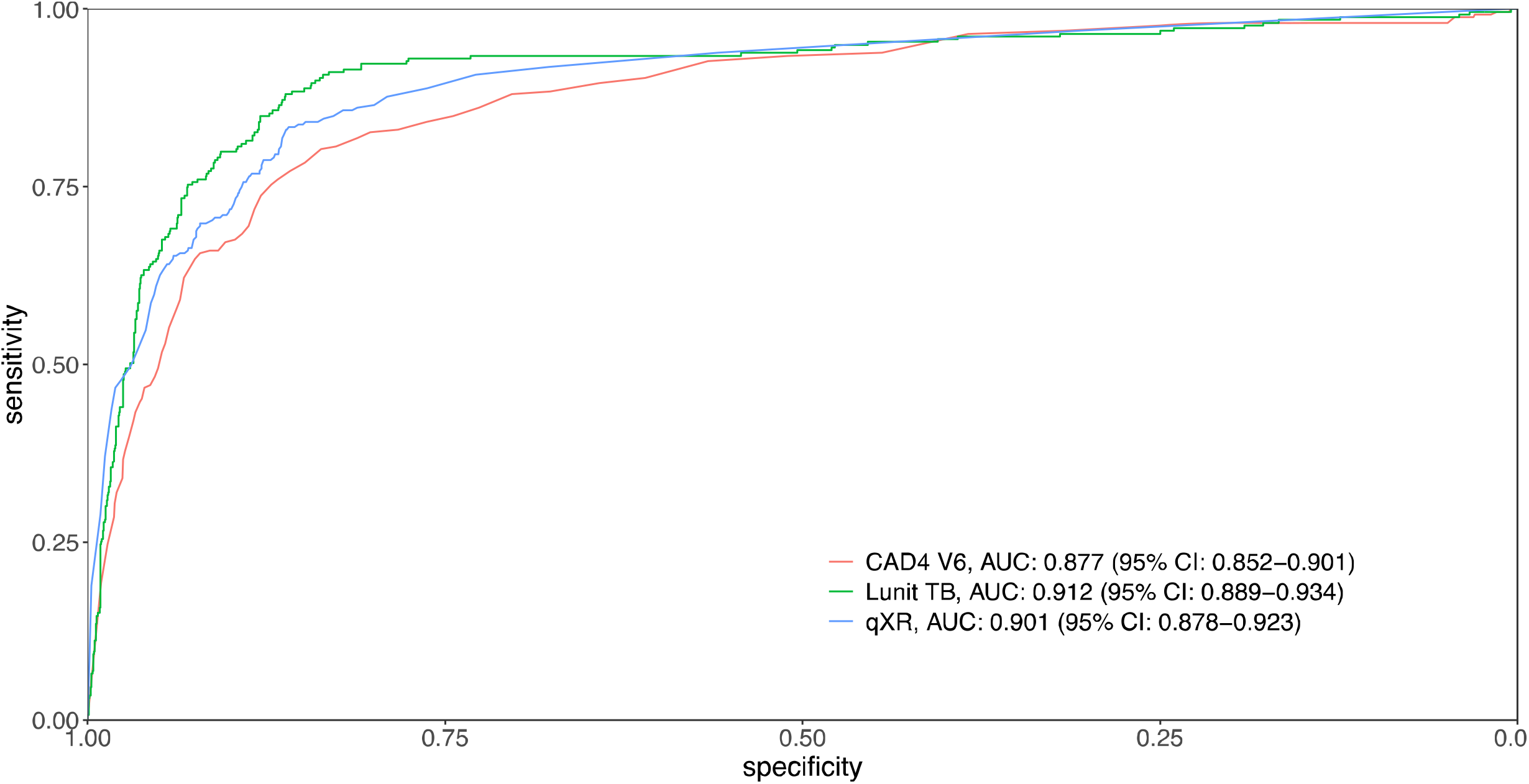
Receiver operating characteristic (ROC) curves for CAD4v6, LunitTB and qXR.

We next performed multivariable logistic regression analysis to examine whether the performance of each algorithm varied by sociodemographic characteristics and risk factors, namely: age, race/ethnicity, current smoker, drug use, previous TB, and TB symptoms.

Specificity of all three algorithms decreased with age and tended to be lower among current smokers and those without TB symptoms, compared to their respective counterparts (**Figure 2, Figure E2-E3)**. LunitTB was the only algorithm that met WHO TPP criteria among individuals 45 years and older. Notably, specificity was under 50% across all three algorithms for individuals with a history of TB.

**Figure 2:**
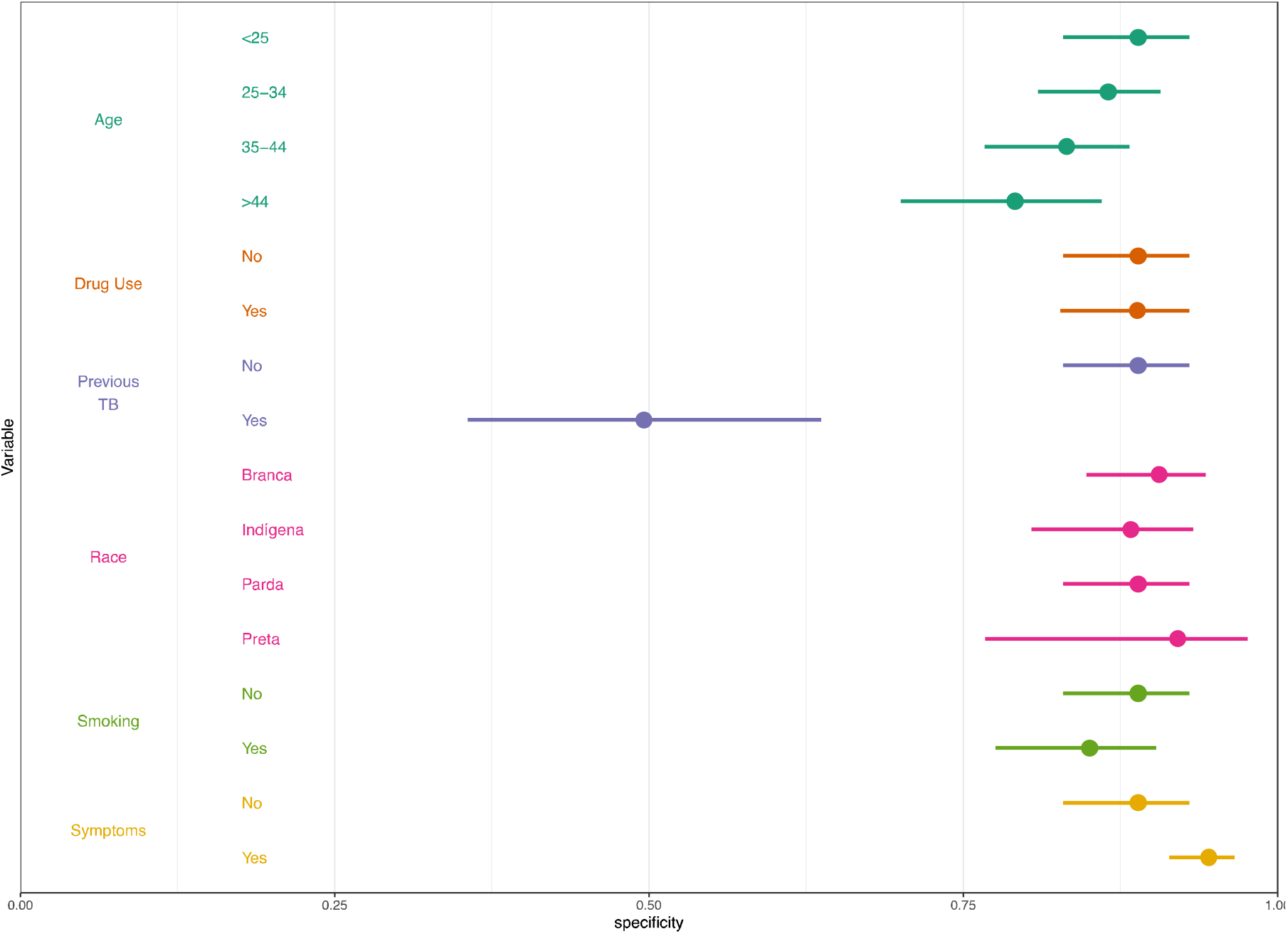
Specificity of LunitTB, by sociodemographic characteristics and risk factors. Shown are the predicted margins for specificity and 95% confidence intervals from a multivariable logistic regression, holding sensitivity at 90%.

To further investigate diagnostic performance depending on history of previous TB, we analyzed the distribution of abnormality scores for TB cases versus non-TB cases as confirmed by sputum Xpert or culture, disaggregated by history of TB. We focused on LunitTB for this analysis given its superior overall performance and its relatively stable specificity by subgroup compared to the other two algorithms. Strikingly, the thresholds required to reach WHO TPP benchmarks of 90% sensitivity and 70% specificity varied dramatically by history of TB (**Figure 3)**. In participants without previous TB, LunitTB score thresholds ≥0.04 had 70% specificity and those ≤0.15 had 90% sensitivity, providing a range of thresholds (0.04-0.15) meeting TPP benchmarks. Conversely, in participants with previous TB, a threshold of at least 0.73 was required for 70% specificity, and there was no score threshold to satisfy both TPP sensitivity and specificity. In participants with previous TB, neither CAD4TBv6 nor qXR had a score that satisfies both TPP sensitivity and specificity. (Figure S4-S5).

**Figure 3:**
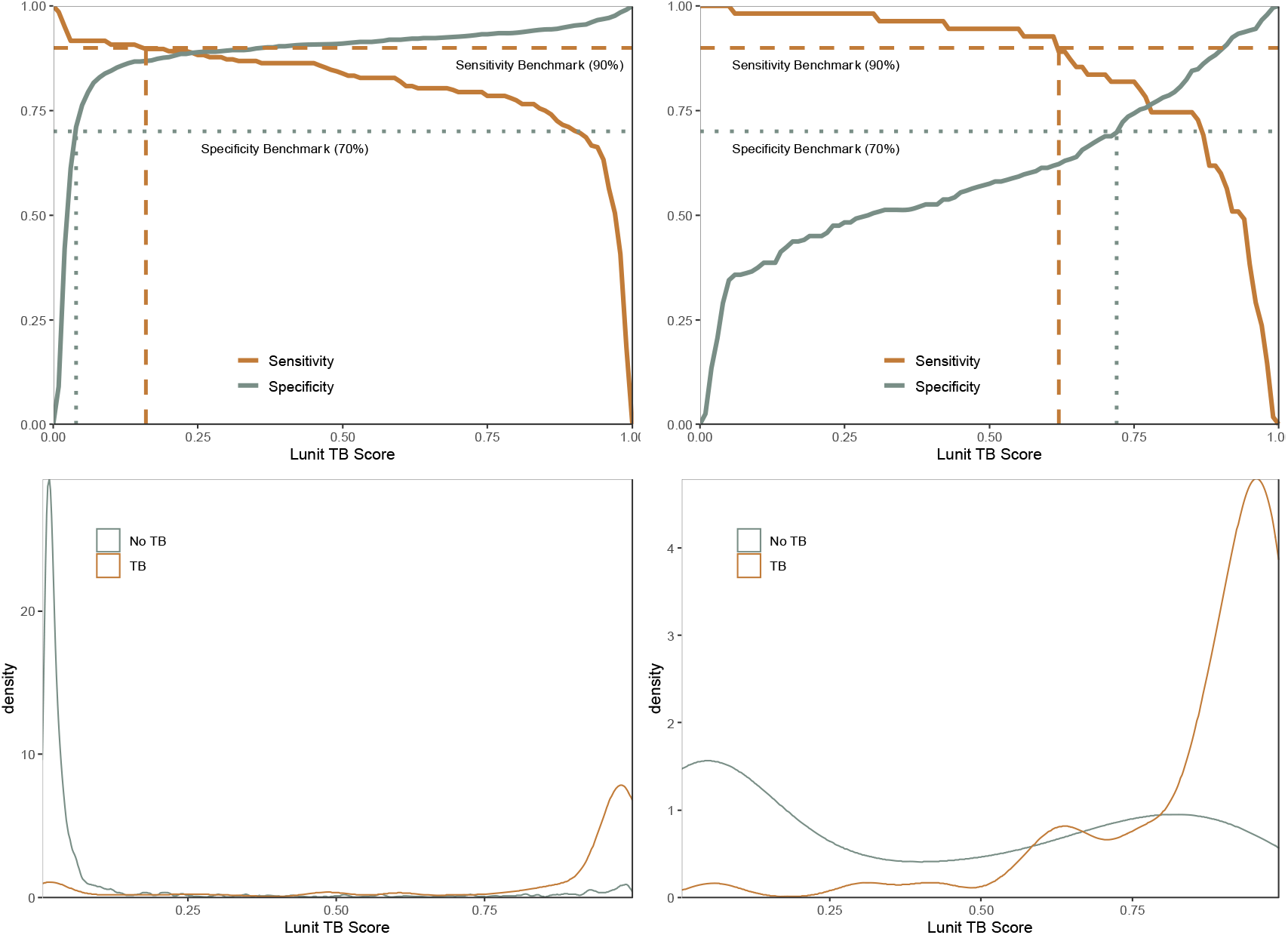
Sensitivity and specificity according to LunitTB score threshold, with the WHO sensitivity (dashed line) and specificity (dotted line) benchmarks (top) among individuals without (A) and with (B) previous TB. Distribution of LunitTB scores for participants without (C) or with (D) previous TB (bottom).

Finally, we assessed the relationship between sputum bacillary load and algorithm performance by examining x-ray abnormality scores by Xpert semi-quantitative result (negative, very low, low, medium, high). Among TB cases with a positive Xpert test for whom x-ray scores were available (188/242), all three algorithms yielded abnormality scores that were positively correlated with sputum Xpert semi-quantitative levels (p<0.0001) **(Figure 4)**. Among the 67 participants with a medium or high Xpert result, CAD4TBv6 had 97% sensitivity (65/67) and LunitTB and qXR both had 96% sensitivity (64/67) at the 70% specificity threshold (**Table E2**).

**Figure 4.**
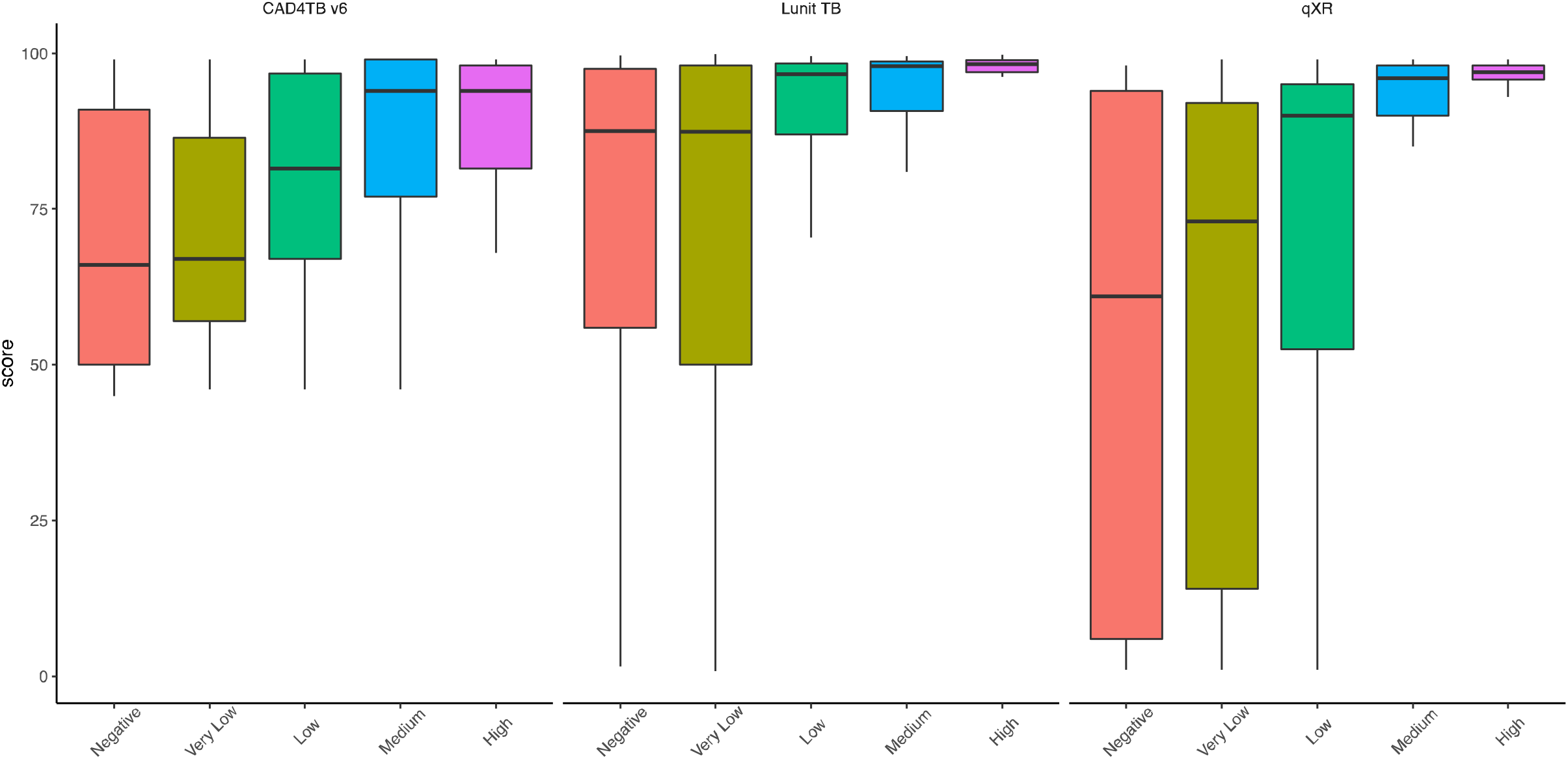
Relationship between the bacillary load in the sputum and the performance of the algorithm through the stratification of the scores by the semiquantitative Xpert result.

## DISCUSSION

Active case finding for tuberculosis in high burden carceral settings is needed to address the substantial excess burden among incarcerated populations. However, despite WHO recommendations for routine TB screening in prisons, most facilities in low- and middle-income countries do not perform systematic active case finding, often citing resource and infrastructure constraints. Effective, cost-efficient screening strategies are needed to make active case finding more accessible in such environments. In this study, conducted via a nurse-led mobile diagnostic unit in three prisons in Central-Western Brazil, we found a very high prevalence of undiagnosed, microbiologically confirmed TB (3.7%). Algorithms for automated interpretation of x-rays achieved high sensitivity and specificity as a screening tool, with the LunitTB and qXR systems exceeding the WHO minimal TPP thresholds for a triage test. Sputum molecular testing is still needed to confirm TB, but a limiting factor in the speed and costs of screening has been the number of tests that can be run daily during mass screening of thousands of individuals. Our findings suggest that screening by mobile x-ray systems with automated interpretation could reduce the number of confirmatory tests required and enable screening to be more rapid and cost-effective in high burden TB settings, while still maintaining sufficient sensitivity.

Recent studies have evaluated x-ray interpretation algorithms among individuals presenting to clinics with TB symptoms, finding variable results. An individual participant meta-analysis found that none of the systems investigated met the WHO TPP criteria for triage, with specificities ranging from 54-61% at 90% sensitivity (14). By contrast, a study in Bangladesh found that the qXR system achieved 74% specificity at the same threshold, and that all algorithms outperformed interpretation by radiologists (15). Our study differed in that it was performed in the context of active case finding, irrespective of symptoms, which could affect estimates of diagnostic accuracy in several ways. For instance, the cases identified through systematic screening are often those in early stages of disease, with lower bacillary burden, as evidenced by the fact that 54% of confirmed cases in our cohort had low, very low, or negative Xpert results. Given the association we observed between sputum bacillary load and x-ray scores, this could have resulted in the algorithms having lower sensitivity in our cohort. At the same time, we might expect higher specificity in the context of active case finding, regardless of symptoms, compared to use in clinics among those presenting with TB symptoms, as the latter setting may include more patients with other pulmonary diseases such as bacterial and viral pneumonias that can be challenging to distinguish from TB. Furthermore, we evaluated these algorithms in incarcerated populations, which tend to be younger, predominantly male, and with high prevalence of various risk factors for TB.

LunitTB was the best-performing algorithm in this cohort, with greater accuracy and generalizability among subgroups, with particularly superior robustness to age compared to the other two algorithms. Nonetheless, performance of all three algorithms varied by subgroup, with consistently lower specificity among older individuals and those with previous TB, corroborating previous findings (15, 16). We also found reduced specificity among current smokers and those without TB symptoms. Therefore, different thresholds may be considered for individuals with different demographic or clinical characteristics. Of note, our pre-defined thresholds for each algorithm led to overall sensitivity under 90%, suggesting that setting-or population-specific threshold calibration may be an important step in implementation.

Specificity of all three algorithms decreased considerably to less than 50% for those with previous TB, indicating failure to meet the WHO TPP for this subgroup. For the 3 analyzed software tools, the distribution of abnormality scores among non-TB cases was shifted higher for those with a history of TB, suggesting the algorithm may not distinguish active TB lesions from fibrous scarring of the lung parenchyma and other chest radiograph patterns indicative of previous TB (17). Thus, in populations with high prevalence of previous TB, Xpert may be more appropriate for screening (18).

We found that x-ray scores were higher—suggestive of more abnormalities—in individuals with high sputum bacillary loads. At the 70% specificity threshold, sensitivity for individuals with medium or high bacillary loads exceeded 96% for all three systems. Given that Xpert bacillary load correlates with smear status (19), and smear status predicts infectiousness (20, 21), it may be reasonable to infer that x-ray automated interpretation algorithms may be more sensitive in identifying the most infectious individuals.

Even with the availability of automated interpretation algorithms, the cost-effectiveness of using x-rays for mass screening in prisons is still unclear. Previous work found that mass screening in prisons with sputum Xpert alone had high yield and was less costly than using x-ray and CAD4TB (version 5) for triage prior to confirmatory Xpert (19). However, the prior study used a single CAD4TB threshold for all individuals and evaluated an additional strategy where only individuals without symptoms were screened with x-ray and CAD4TB prior to confirmatory Xpert. Our present findings suggest that such strategies may be less effective due to the algorithms’ variable performance by subgroup, particularly the reduced sensitivity specificity for individuals without TB symptoms. Furthermore, CAD4TB (version 6) was shown to have the lowest performance in this study; thus, screening with a more accurate algorithm like LunitTB could increase cost-effectiveness. Additionally, emerging technologies for portable, digital radiography could reduce consumable costs, making x-rays more accessible and affordable in resource-constrained environments.

This study has several limitations. First, in our primary analysis, we only included participants who were able to produce sputum for confirmatory testing, as sputum induction was not able to be undertaken in this environment. The excluded participants were less likely to be current smokers, to have TB symptoms, and to report previous TB; we expect that their inclusion may have affected overall estimates of algorithm performance in this population. In secondary analyses of the entire population, AUCs did not differ significantly. However, future research is needed to evaluate these x-ray interpretation algorithms on this group, given that a strength of x-ray screening is the lack of requirement for sputum. Second, we used solid media culture due to local availability costs; however, solid media culture is less sensitive than liquid media and could have led to missed cases. Additionally, we note that while for LunitTB and qXR we used the manufacturers’ recommended thresholds, for CAD4TBv6 we used a threshold determined from a subset of our population; therefore, the thresholds at 90% sensitivity may be more appropriate than our pre-defined thresholds for comparison of the three algorithms. Due to the low prevalence of HIV in our population, we did not consider HIV status in our study (22). Moreover, our study only included those in male prisons as there are fewer than 10 cases annually among incarcerated women in this state; consequently, the performance of these algorithms for TB screening in female prisons remains unknown (16).

Overall, our results suggest that the use of chest x-rays and artificial intelligence-based interpretation algorithms can be part of an effective mass screening strategy in high-burden settings like prisons. Although Lunit-TB had the greatest accuracy and robustness in our cohort, all three algorithms exhibited similar performance, particularly as a rule-out-test, and could be used to reduce the need for universal molecular testing. However, our findings suggest the need for future optimization of these algorithms to improve generalizability across subgroups, especially for individuals with a history of TB. Nevertheless, given their high overall accuracy in this population, especially among cases with the greatest sputum bacillary load, automated interpretation algorithms could enable scaling of mass screening to help mitigate disparities in TB diagnosis among incarcerated populations.

## Data Availability

All data produced in the present work are contained in the manuscript.

## ACKNOWLEDGMENTS

The State Agency for the Administration of the Penitentiary System of the State of Mato Grosso do Sul (AGEPEN) for having authorized the study; the Coordination for the Improvement of Higher Education Personnel (CAPES); the Federal University of Grande Dourados (UFGD) for having supported the study in their postgraduate programs and the Qure.ai Technologies Private Limited for the x-ray images analysis.

## DATA SUPPLEMENT – Tables and Figures

**Table E1.** Sociodemographic characteristics and risk factors for TB by study inclusion/exclusion. ExcludE refers to those excludE due to lack of a confirmatory sputum Xpert or culture result.

| **Variables** | **Included**<br>**N=2075 (%)** | **Excluded**<br>**N=4940 (%)** | **P Value** |
| --- | --- | --- | --- |
| Median age, (IQR) | 33 (28, 40) | 34 (28, 40) | <0.001 |
| Prison Unit |  |  |  |
| PED | 889 (42.8) | 1444 (29.2) | <0.001 |
| EPJFC | 840 (40.5) | 1940 (39.3) |  |
| IPCG | 346 (16.9) | 1556 (31.4) |  |
| Ethnic/skin color |  |  |  |
| Mixed | 1279 (61.6) | 3068 (62.1) | 0.428 |
| White | 508 (24.5) | 1115 (22.6) |  |
| Black | 253 (12.2) | 612 (12.4) |  |
| Indigenous | 33 (1.6) | 138 (2.8) |  |
| Asian | 2 (0.1) | 7 (0.1) |  |
| <8y of schooling | 1546 (74.5) | 3511 (71.1) | 0.003 |
| Current smoker | 1520 (73.3) | 2737 (55.4) | <0.001 |
| BCG vaccinated | 1862 (89.7) | 4378 (88.6) | 0.197 |
| Previous TB | 293 (14.1) | 268 (5.4) | <0.001 |
| Report any WHO TB symptoms | 1512 (72.9) | 851 (17.2) | <0.001 |
| Report cough | 1255 (60.5) | 536 (10.9) | <0.001 |
| Illicit drug use over the last year | 1460 (70.4) | 2644 (53.5) | <0.001 |
| Previously incarcerated | 1557 (75.0) | 3478 (70.4) | <0.001 |
| TB contact | 1565 (75.4) | 3125 (63.3) | <0.001 |

**Abbreviations:** BCG: Bacillus of Calmette and Guérin; IQR: interquartile range; TB: tuberculosis; WHO: World Health Organization, y: years

**Table E2.** Sensitivity of x-ray interpretation algorithms according to Xpert MTB/RIF G4 semi-quantitative load, using a threshold for specificity of 70%.

| Sputum Xpert result | n | CAD4TBv6 |  | LunitTB |  | qXR |  |
| --- | --- | --- | --- | --- | --- | --- | --- |
|  |  | Positive | Sens. (95% CI) | Positive | % Sens. (95% CI) | Positive | % Sens. (95% CI) |
| Negative/Very Low | 60 | 47 | 78.3 (65.8-87.9) | 53 | 88.3 (77.4-95.1) | 49 | 81.7 (69.6-90.5) |
| Low | 78 | 70 | 89.7 (80.8-95.5) | 73 | 93.6 (85.7-97.9) | 73 | 93.6 (85.7-97.9) |
| Medium/High | 67 | 65 | 97.0 (89.6-99.6) | 64 | 95.5 (87.5-99.1) | 64 | 95.5 (87.5-99.1) |

**Figure E1.**
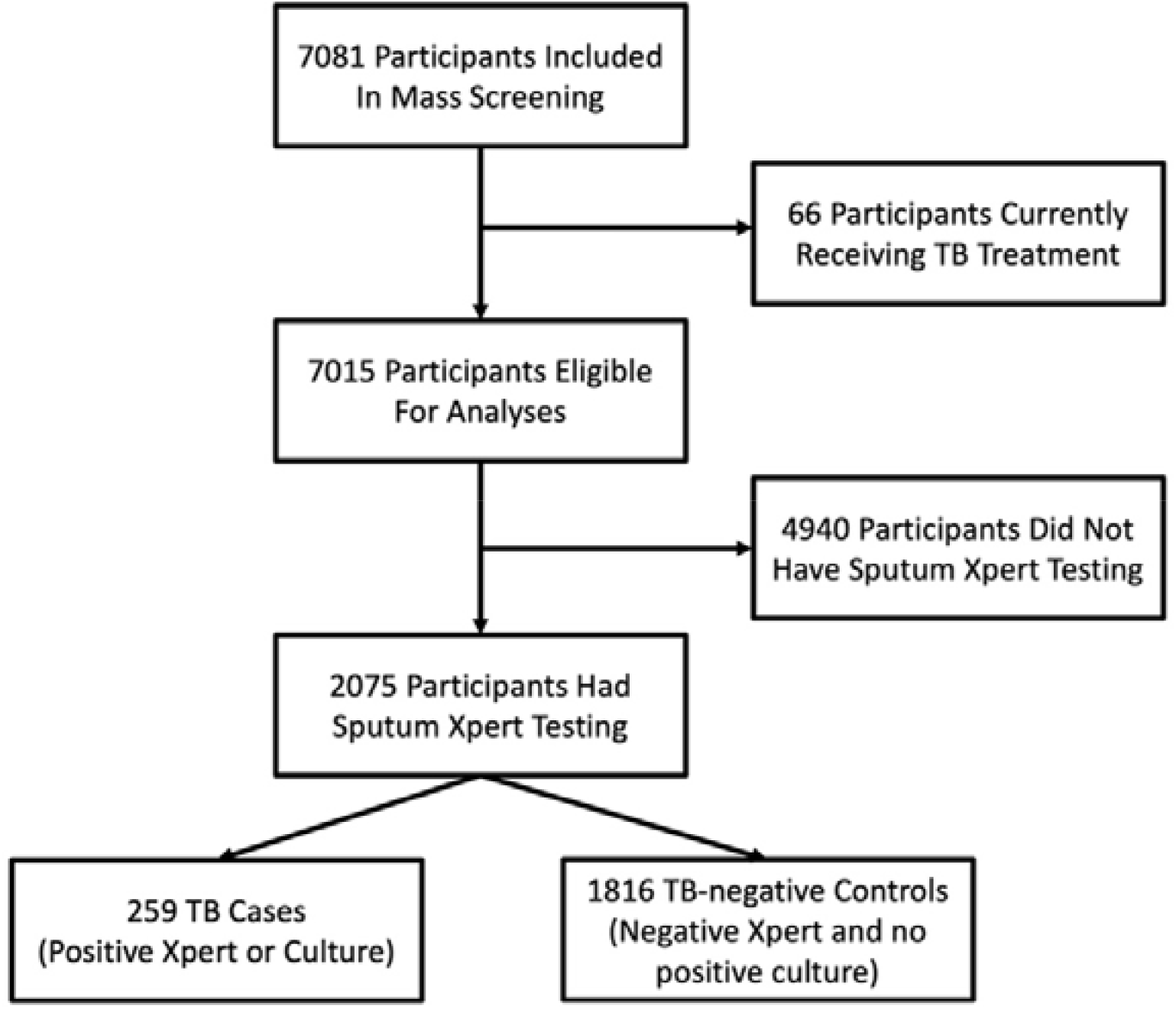
Flow diagram of study participants in mass screening and inclusion of participants in x-ray evaluation.

**Figure E2:**
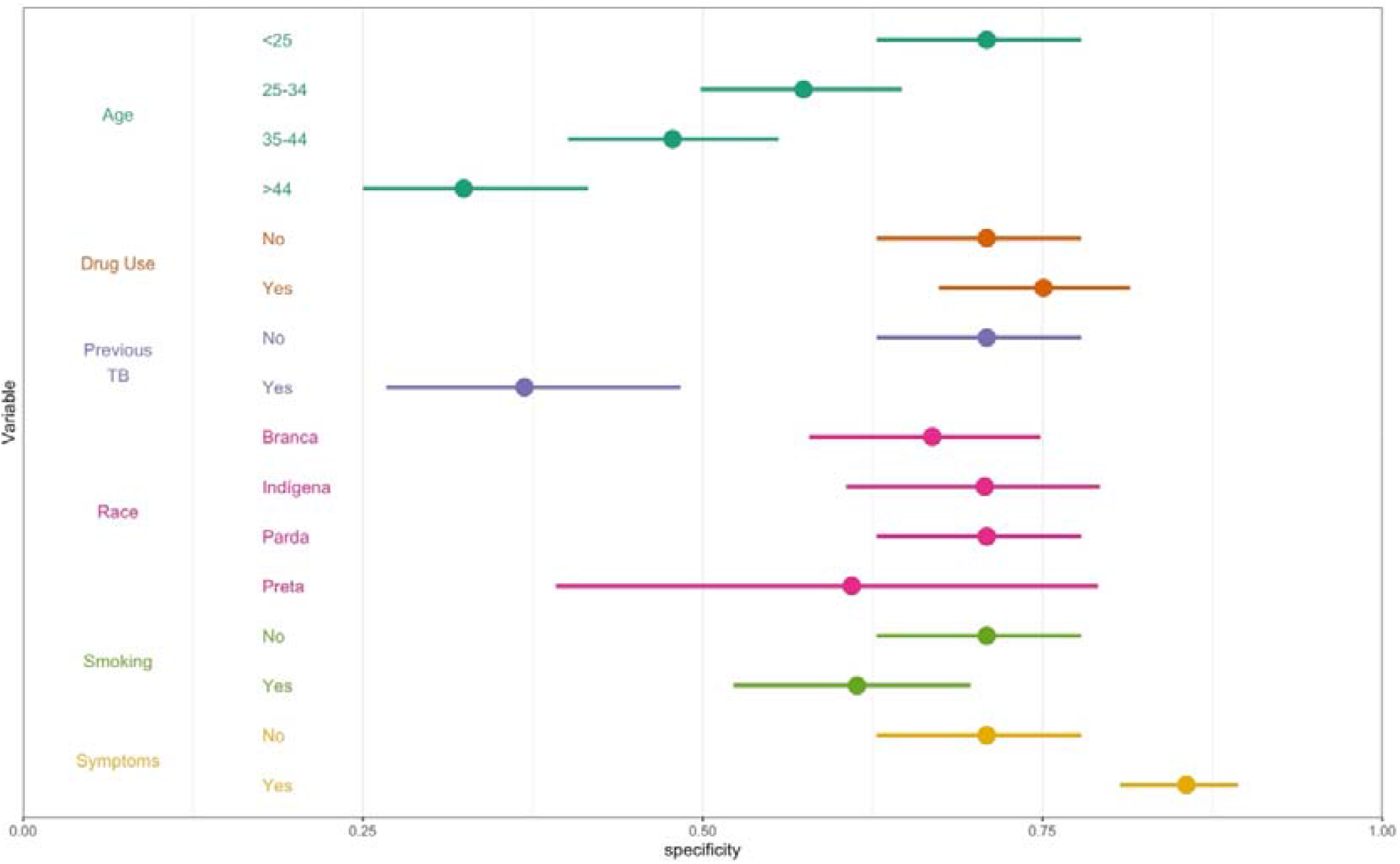
Specificity of CAD4TBv6, by sociodemographic characteristics and risk factors. Shown are the predicted margins for specificity and 95% confidence intervals from a multivariable logistic regression, holding sensitivity at 90%.

**Figure E3:**
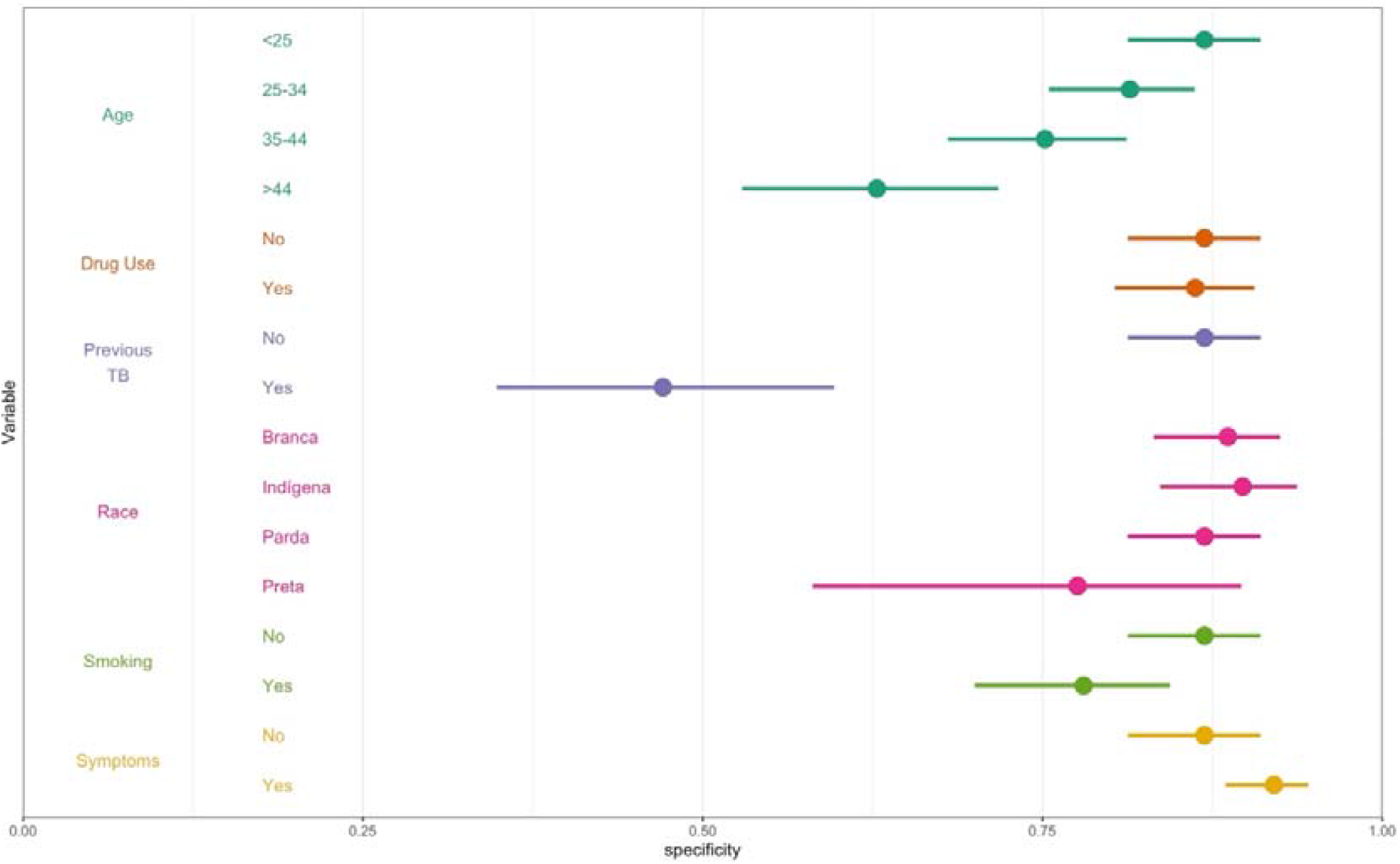
Specificity of qXR, by sociodemographic characteristics and risk factors. Shown are the predicted margins for specificity and 95% confidence intervals from a multivariable logistic regression, holding sensitivity at 90%.

**Figure E4:**
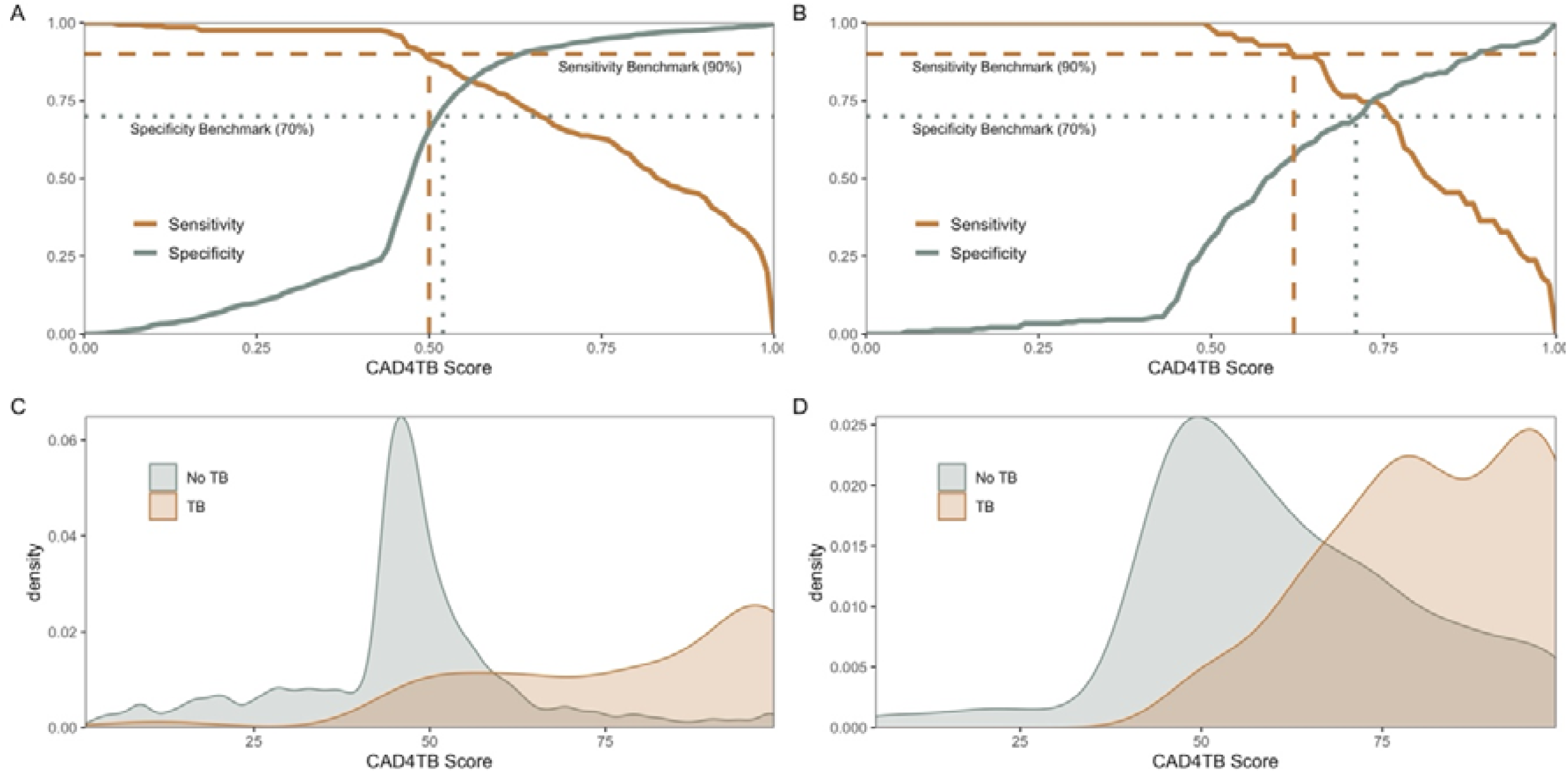
Sensitivity and specificity according to CAD4TBv6 score threshold, with the WHO sensitivity (dashE line) and specificity (dottE line) benchmarks (top) among individuals without (A) and with (B) previous TB. Distribution of CAD4TBv6scores for participants without (C) or with (D) previous TB (bottom).

**Figure E5:**
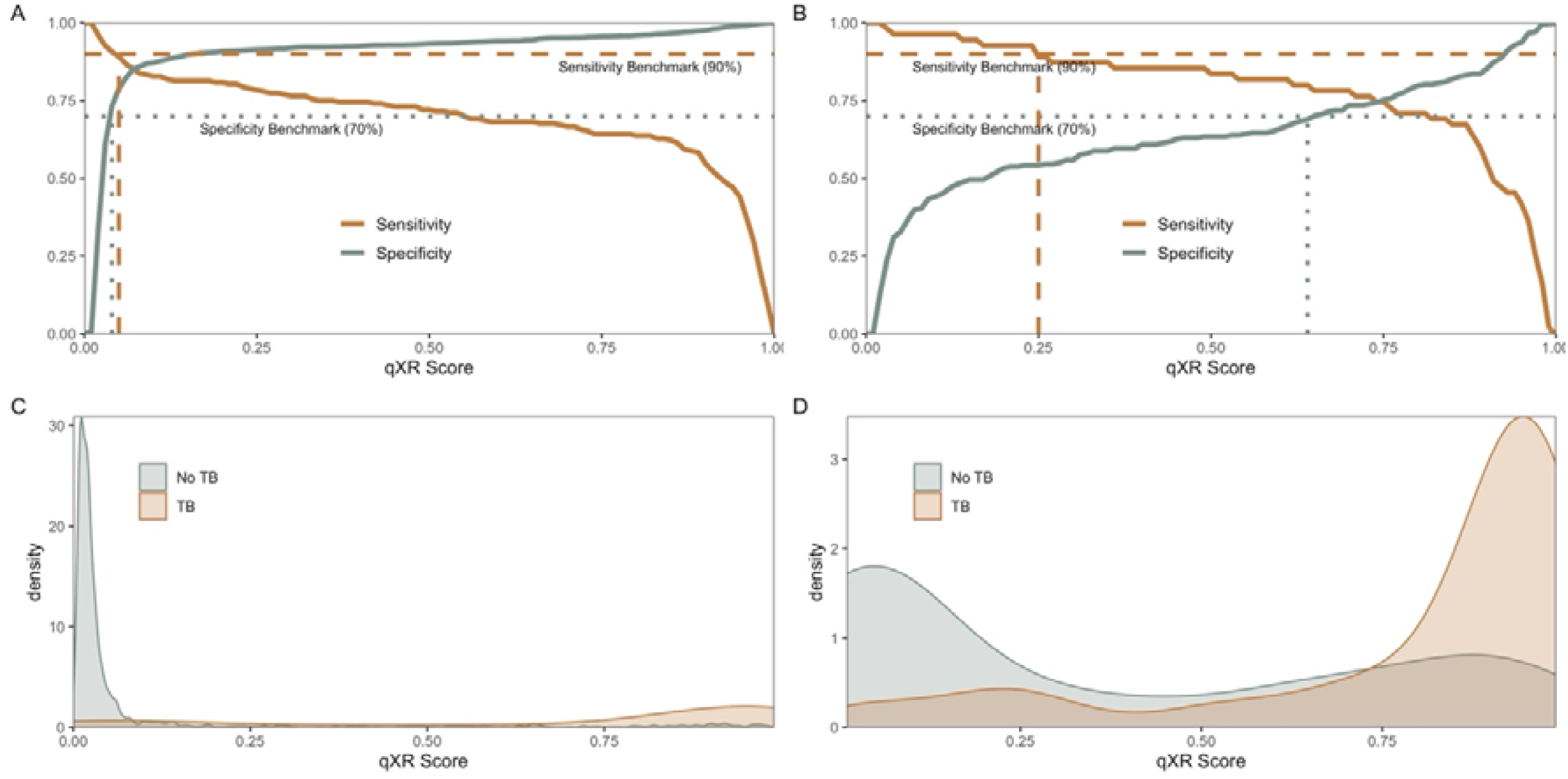
Sensitivity and specificity according to qXR score threshold, with the WHO sensitivity (dashE line) and specificity (dottE line) benchmarks (top) among individuals without (A) and with (B) previous TB. Distribution of qXR scores for participants without (C) or with (D) previous TB (bottom).

## Notes

**Sources of support:** This study was supported by the US National Institutes of Health (grant numbers R01 AI130058 and R01 AI149620) and the State Secretary of Health of Mato Grosso do Sul.

### Competing Interest Statement

The authors have declared no competing interest.

### Funding Statement

This study was funded by US National Institutes of Health (grant numbers R01 AI130058 and R01 AI149620) and the State Office of Health of Mato Grosso of Sul.

### Author Declarations

Ethics committee/IRB of Federal University of Grande Dourados and Stanford University gave ethical approval for this work.

## REFERENCES

1. Cords O, Martinez L, Warren JL, O’Marr JM, Walter KS, Cohen T, Zheng J, Ko AI, Croda J, Andrews JR. Incidence and prevalence of tuberculosis in incarcerated populations: a systematic review and meta-analysis. Lancet Public Health 2021;6:e300–e308.

2. Walter KS, Martinez L, Arakaki-Sanchez D, Sequera VG, Sanabria GE, Cohen T, Ko AI, García-Basteiro Al, Rueda ZV, López-Olarte Ra, Espinal MA, Croda J, Andrews JR. The escalating tuberculosis crisis in central and South American prisons. The Lancet 2021;397:1591–1596.

3. World Health Organization. WHO consolidated guidelines on tuberculosis. Module 2: screening – systematic screening for tuberculosis disease. Geneva: World Health Organization; 2021.

4. Hermans SM, Andrews JR, Bekker L-G, Wood R. The mass miniature chest radiography programme in Cape Town, South Africa, 1948 - 1994: The impact of active tuberculosis case finding. South Afr Med J Suid-Afr Tydskr Vir Geneeskd 2016;106:1263–1269.

5. Comstock GW, Philip RN. Decline of the tuberculosis epidemic in Alaska. Public Health Rep Wash DC 1896 1961;76:19–24.

6. Organization WH. WHO Expert Committee on Tuberculosis [meeting held in Geneva from 11 to 20 December 1973]: ninth report. 1974;

7. World Health Organization. Systematic screening for active tuberculosis: an operational guide. 2015. At <http://apps.who.int/iris/bitstream/10665/181164/1/9789241549172_eng.pdf?ua=1>.

8. Pinto LM, Pai M, Dheda K, Schwartzman K, Menzies D, Steingart KR. Scoring systems using chest radiographic features for the diagnosis of pulmonary tuberculosis in adults: a systematic review. Eur Respir J 2013;42:480–494.

9. Harris PA, Taylor R, Minor BL, Elliott V, Fernandez M, O’Neal L, McLeod L, Delacqua G, Delacqua F, Kirby J, Duda SN. The REDCap consortium: Building an international community of software platform partners. J Biomed Inform 2019;95:103208.

10. Harris PA, Taylor R, Thielke R, Payne J, Gonzalez N, Conde JG. Research electronic data capture (REDCap)—A metadata-driven methodology and workflow process for providing translational research informatics support. J Biomed Inform 2009;42:377–381.

11. Nash M, Kadavigere R, Andrade J, Sukumar CA, Chawla K, Shenoy VP, Pande T, Huddart S, Pai M, Saravu K. Deep learning, computer-aided radiography reading for tuberculosis: a diagnostic accuracy study from a tertiary hospital in India. Sci Rep 2020;10:210.

12. World Health Organization. High-priority target product profi les for new tuberculosis diagnostics: report of a consensus meeting. World Health Organization; 2014. At <http://apps.who.int/iris/bitstream/handle/10665/135617/WHO_HTM_TB_2014.18_eng.pdf;jsessionid=0CBFF2089B61E11AFF4C116E2CE6E273?sequence=1>.

13. pROC: an open-source package for R and S+ to analyze and compare ROC curves. At <https://bmcbioinformatics.biomedcentral.com/articles/10.1186/1471-2105-12-77>.

14. Tavaziva G, Harris M, Abidi S, Geric C, Breuninger M, Dheda K, Esmail A, Muyoyeta M, Reither K, Majidulla A, Khan A, Campbell J, David P-M, Denkinger C, Miller C, Nathavitharana R, Pai M, Benedetti A, Ahmad Khan F. Chest X-ray Analysis With Deep Learning-Based Software as a Triage Test for Pulmonary Tuberculosis: An Individual Patient Data Meta-Analysis of Diagnostic Accuracy. Clin Infect Dis 2021;doi:10.1093/cid/ciab639.

15. Qin ZZ, Ahmed S, Sarker MS, Paul K, Adel ASS, Naheyan T, Barrett R, Banu S, Creswell J. Tuberculosis detection from chest x-rays for triaging in a high tuberculosis-burden setting: an evaluation of five artificial intelligence algorithms. Lancet Digit Health 2021;3:e543–e554.

16. Khan FA, Majidulla A, Tavaziva G, Nazish A, Abidi SK, Benedetti A, Menzies D, Johnston JC, Khan AJ, Saeed S. Chest x-ray analysis with deep learning-based software as a triage test for pulmonary tuberculosis: a prospective study of diagnostic accuracy for culture-confirmed disease. Lancet Digit Health 2020;2:e573–e581.

17. Piccazzo R, Paparo F, Garlaschi G. Diagnostic accuracy of chest radiography for the diagnosis of tuberculosis (TB) and its role in the detection of latent TB infection: a systematic review. J Rheumatol Suppl 2014;91:32–40.

18. Frascella B, Richards AS, Sossen B, Emery JC, Odone A, Law I, Onozaki I, Esmail H, Houben Rmgj. Subclinical tuberculosis disease - a review and analysis of prevalence surveys to inform definitions, burden, associations and screening methodology. Clin Infect Dis doi:10.1093/cid/ciaa1402.

19. Beynon F, Theron G, Respeito D, Mambuque E, Saavedra B, Bulo H, Sanz S, Dheda K, Garcia-Basteiro AL. Correlation of Xpert MTB/RIF with measures to assess Mycobacterium tuberculosis bacillary burden in high HIV burden areas of Southern Africa. Sci Rep 2018;8:5201.

20. Behr MA, Warren SA, Salamon H, Hopewell PC, Ponce de Leon A, Daley CL, Small PM. Transmission of Mycobacterium tuberculosis from patients smear-negative for acid-fast bacilli. Lancet Lond Engl 1999;353:444–449.

21. Hernández-Garduño E, Cook V, Kunimoto D, Elwood RK, Black WA, FitzGerald JM. Transmission of tuberculosis from smear negative patients: a molecular epidemiology study. Thorax 2004;59:286–290.

22. Sgarbi RVE, Carbone A da SS, Paião DSG, Lemos EF, Simionatto S, Puga MAM, Motta-Castro ARC, Pompilio MA, Urrego J, Ko AI, Andrews JR, Croda J. A Cross-Sectional Survey of HIV Testing and Prevalence in Twelve Brazilian Correctional Facilities. PloS One 2015;10:e0139487.

